# Validation of Natural Language Processing for Surgical Complication Surveillance: Detecting Eleven Postoperative Complications from Electronic Health Records

**DOI:** 10.1101/2025.04.07.25325367

**Authors:** Emilie E. Dencker, Alexander Bonde, Anders Troelsen, Martin Sillesen

## Abstract

**Background:** Postoperative complications (PCs) rates are crucial quality metrics in surgery, as they reflect both patient outcomes, perioperative care effectiveness and healthcare resource strain. Despite their importance, efficient, accurate, and affordable methods for tracking PCs are lacking. This study aimed to evaluate whether natural language processing (NLP) models could detect eleven PCs from surgical electronic health records (EHRs) at a level comparable to human curation.

**Methods:** 17 486 surgical cases from 18 hospitals across two regions in Denmark, spanning six years, were included. The dataset was divided into training, validation, and test sets for NLP-model development and evaluation (50.2%/33.6%/16.2%). Model performance was compared against the current method of PC monitoring (ICD-10 codes) and manual curation, the latter serving as the gold standard.

**Results:** The NLP-models had a ROC AUC between 0.901 to 0.999 for the test set. Sensitivity of the models when compared to manual curation ranged from 0.701 to 1.00, except for myocardial infarction (0.500). Positive Predictive Value (PPV) ranged from 0.0165 to 0.947, and Negative Predictive Value from 0.995 to 1.00. The NLP-models significantly outperformed ICD-10 coding in detecting PC, resulting in 16.3% of cases would require manual curation to reach a PPV of 1.00

**Conclusion:** The NLP models alone were able to detect PCs at an acceptable level and performed superior to ICD-10 codes. Combining NLP based and manual curation was required to reach a PPV of 1.00. Therefore, NLP algorithms present a potential solution for comprehensive and real-time monitoring of PCs across the surgical field.

## INTRODUCTION

Postoperative complications (PCs) are widely recognized as a critical measure of surgical quality(1).

PCs reflect not only the surgical technical performance but also the quality of the peri- and postoperative care, impacting patient outcomes, including quality of life, functional level, and life expectancy. Additionally, at a systemic level, PCs demand substantial resources, both in terms of clinical staffing and economic costs(2–5).

Despite their importance, methods for detecting and monitoring PCs remain debated. Many healthcare systems use administrative data sources, such as ICD-10 codes and insurance claims, to track PC rates. However, these data sources are often criticized for limited reliability(6–10), with the most notable issues being underreporting and inaccurate documentation of PCs(5,11–13).

To overcome these limitations, some healthcare systems have developed quality improvement programs relying on manual data extraction from electronic healthcare records (EHRs). The American College of Surgeons’ National Surgical Quality Improvement Program (ACS NSQIP) being a prominent example. While effective and cost-efficient(14), these programs have limitations: they are resource-intensive process, can introduce selection bias, and lack real-time monitoring capacity(15,16).

Considering the limitations of the current monitoring methods and the resource strain associated with PCs, there is an urgent need for a real-time, high-quality monitoring approach. Here, artificial intelligence (AI), particularly natural language processing (NLP), offers a promising opportunity to improve surgical quality by analyzing text from EHRs with minimal cost and time(6,18–20). Yet, the range of PCs that NLP can accurately detect, remains unclear.

This study addresses this gap by evaluating NLP algorithms for detecting a wide spectrum of PCs and assessing their potential as a universal monitoring tool across various surgical specialties.

Specifically, we aimed to determine whether NLP models could identify eleven target PCs (deep surgical site infection, organ/space site infection, wound disruption, pulmonary embolism, stroke, cardiac arrest, myocardial infarction, deep vein thrombosis, unplanned readmission, unplanned reoperation, and unplanned intubation) from EHR data, including free medical text, with accuracy comparable to human curation. We hypothesized that NLP-models would reliably detect these PCs up to 30 days postoperatively, achieving performance levels equivalent to human curation.

## METHODS

The study received approval from the Institutional Review Board (IRB) for retrospective patient studies in Denmark, the Danish Patients Safety Board (Styrelsen for Patientsikkerhed, approval #31-1521-182), and the Danish Capital Region Data Safety Board (Videnscenter for Dataanmeldelser, approval #P-2020-180). As it was retrospective, utilized de-identified data, and involved no patient contact, informed consent was not required under Danish law.

### Data source

We obtained data from EHRs of surgical patients in Denmark’s Capital and Zealand regions over five years (2016–2021). Denmark operates a tax-funded universal healthcare system providing comprehensive surgical care to all citizens at no personal expense. The two regions encompass 18 hospitals with access to all surgical subspecialities and postsurgical care, serving approximately 2.7 million residents(17). The dataset included all surgical cases within the five-year period, covering a range of surgeries (elective and emergency) across various specialties, including orthopedic, general, plastic, gynecology and obstetrics, urology, ophthalmic, cardiothoracic, vascular, neurological, oral, and otorhinolaryngology surgery.

The EHR data comprised clinical notes (e.g., admission notes, progress notes, procedure notes, discharge notes), biochemical values, and vital signs recorded up to 30 days postoperatively.

Additionally, ICD-10 diagnosis codes for comorbidities, acute conditions and PCs were included along with demographic data (age, sex, weight, etc.).

To prepare data for use in a natural language processing context, the total dataset was split into three separate subsets: a training set, a validation set and a test set (see below). All subsets (training, validation, and test sets) were manually labeled according to the 2018 NSQIP criteria by a team of medical reviewers for the presence of any of the studied complications. The 2018 NSQIP criteria are available in Table S2 in Supplementary Material.

#### Training set

The training set includes cases from May 2016 to October 13st, 2021, and was originally developed for a previous study on superficial surgical site infections (SSSIs). Cases with an ICD-10 code indicating an SSSI were included. (Illustrated in Figure 1 “case flow diagram”). The included cases were manually labeled for various postoperative complications, making the training set suitable for multiple studies on detection of different PCs.

**Figure 1:**
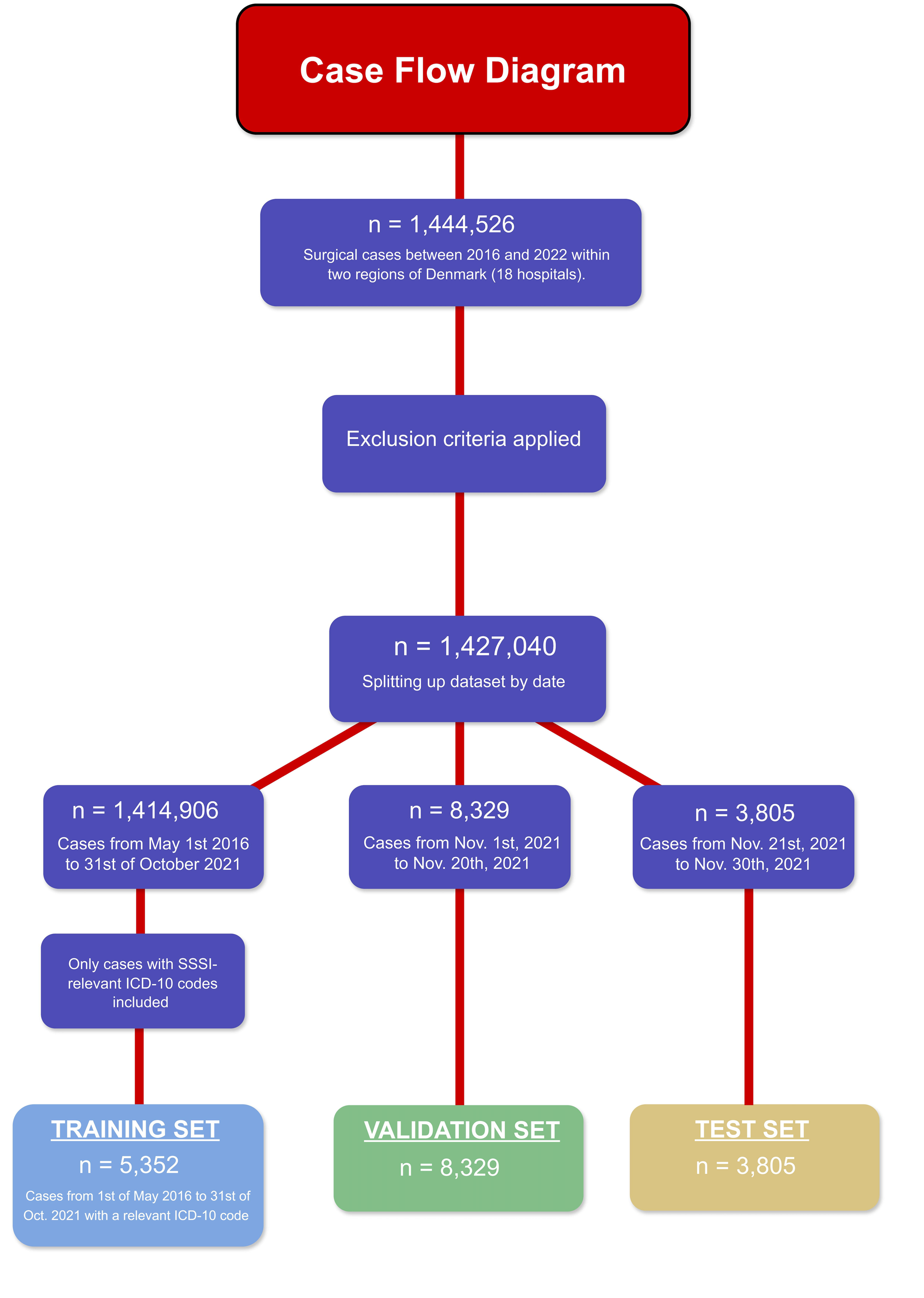
The case flow diagram illustrates the stepwise process of case selection. Cases were excluded based on predefined criteria (see Table S3 in Supplementary Data). After exclusions, the remaining cases were divided into three subsets: the training, validation, and test set, in accordance with the study’s time-based split and inclusion criteria.

When using natural language processing (NLP), the first step is to train the model. This involves using a portion of the data (the training set) for the model to learn how to classify information. This process requires the data being labeled, so that the model learns which features (words or phrases) are important for detecting an outcome. The training data also teaches the model to connect input (medical free text) with output (postoperative complications). Finally, the training data helps the model generalize from specific examples to predictions/classifications on unseen data. After training, classifiers were added – one for each PC. A classifier determines at which probability the given PC being classified as present or not. These first steps of model development are illustrated in Figure 2, step 1.

**Figure 2:**
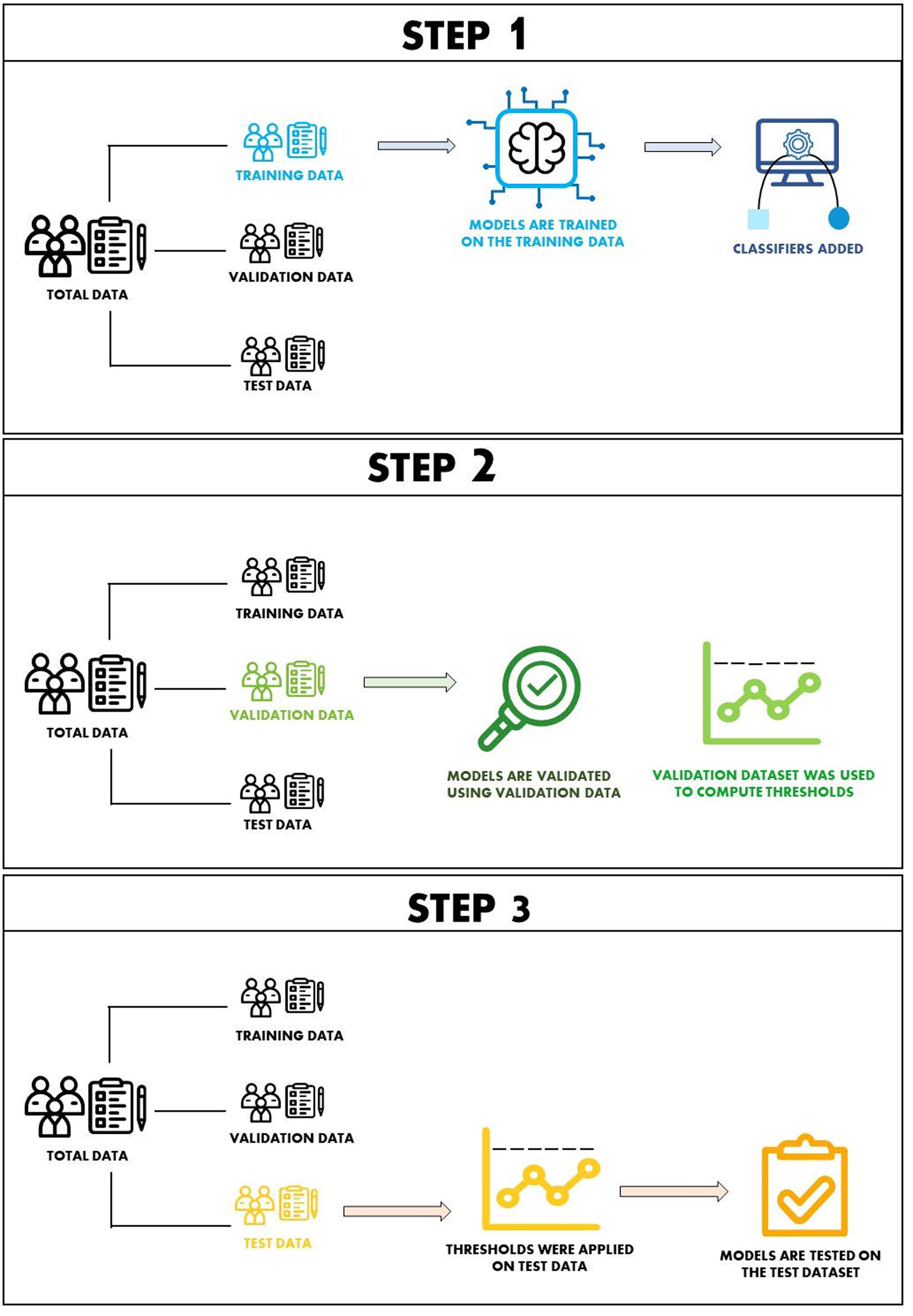
This figure outlines the step-by-step development of the NLP models and the specific roles of the three data subsets. The training set is used to train the models, after which classifiers are added. The validation set is used to validate the models and fine-tune the thresholds. Finally, the test set is used to evaluate the models’ performance, where the previously set thresholds are applied.

#### Validation set

The validation set included cases from November 1^st^ and November 20^th^, 2021 (see Figure 1). The validation set is used to fine-tune model configuration, optimize hyperparameters, and determine thresholds used for classifying PCs (illustrated in Figure 2, step 2). Since classification is expressed as a probability rather than a binary output (yes/no), the threshold defines the probability level at which a PC is flagged as present. Adjusting these thresholds affects model sensitivity and specificity.

#### Test set

The test set covered cases from November 21^st^ and November 30^th^, 2021 (Figure 1). The test set is used to test the model’s performance on a portion of yet unseen data. Therefore, a “ground truth” must be established, and for this we chose manual curation of the EHRs. The thresholds determined during the validation process are then applied to the test set (Figure 2, step 3.)

#### ICD-10 codes

To compare the NLP model’s performance with conventional ICD-10-based monitoring, relevant codes were defined for each PC (Table S1, Supplementary material). ICD-10-codes were documented by treating physicians during treatment or at time of discharge. All codes within 30 days postoperatively were included. For the PCs “unplanned readmission”, “unplanned reoperation”, and “unplanned intubation” no relevant ICD-10 codes exist, hence comparison was limited to NLP-models vs. manual curation.

#### Data leakage and missing data

To reduce data leakage and missing data, several steps were taken. Cases not marked as “complete” and cases with missing values for procedure start or completion times were excluded.

Only cases from relevant surgical specialties (as determined by SKS-code) were included. If a patient had multiple unrelated procedures within 30 days, only the first was included, except in cases of reoperation. To prevent overlap between the subset, notes after October 30^th^, 2021, were excluded from the training set, ensuring no overlap with the validation or test sets.

#### Machine learning modelling

Model training has been detailed previously(6). Briefly, we used a preprocessing approach including Term Frequency-Inverse Document Frequency (TFIDF) with vectorization. Following normalization, the text was forwarded through Light Gradient Boosting Machine Classifiers (LightGBM, Microsoft Azure implementation) serving as the gradient boosting decision tree model.

#### Threshold selection

Since the model outputs probabilities for each PC rather than binary classifications, selecting the optimal threshold for these probabilities is crucial. The thresholds directly affect the sensitivity and specificity of the model, influencing rates of false positives and false negatives. To address this, we assess two approaches: a Stand-Alone-Model (SAM) and a Human-In-The-Loop (HITL). The SAM classifies cases based on probability thresholds alone, while the HITL-approach flags higher- probability cases for human review, while low-probability cases were classified as negative and not reviewed by human. Thresholds were selected to optimize the F2 score, prioritizing sensitivity over positive predictive value, in order to minimize the rate of false negatives.

#### Performance

Model performance was assessed using Receiver Operator Area Under the Curve (ROCAUC), alongside sensitivity, specificity, positive predictive value (PPV), and negative predictive value (NPV). Additionally, thresholds for both the Stand-Alone-Model (SAM) and Human-In-The-Loop (HITL) approach are provided. For comparison, performance metrics for ICD-10 codes are also included.

#### Data presentation and implementation

Results are presented as counts, medians [interquartile range], percentages and performance metrics as stated above where appropriate. Data processing was conducted within a secure cloud environment on the Windows Azure platform. Python v. 3.8.13 was used to implement the NLP algorithms.

## RESULTS

The total dataset comprised 17 486 surgical cases, corresponding to 17 925 surgical procedures and 182 293 EHR chart notes. Data preprocessing steps are outlined in Figure 1 and exclusion criteria are detailed in Table S3 (Supplementary Material). The dataset was split into a training, validation, and test set on a time-based approach (Figure 3). The training set comprised 5 352 cases and 91 435 EHR notes, the validation set comprised 8 329 and 61 335 EHR notes, and the test set comprised 3 805 cases and 29 523 EHR notes. Based on EHR notes the split was 50.2% / 33.6% / 16.2% (training / validation / test).

**Figure 3:**
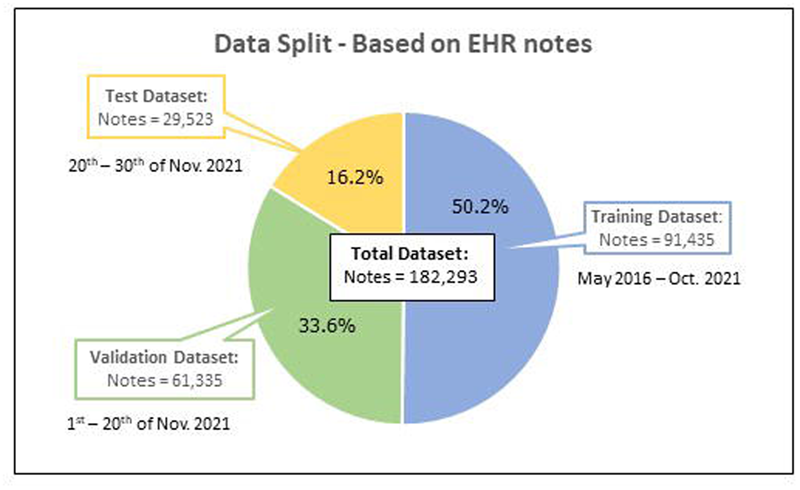
Data split by the number of electronic healthcare record notes, distributed across the training, validation, and test sets using a time-based split.

Demographic results are presented in Table 1. Of the study population, 52.8 % were female, with a median age of 59 years. The median surgery duration was 52 minutes, with the most common surgical specialties being orthopedic (24.4%) and general surgery (22.9%).

**Table 1:**
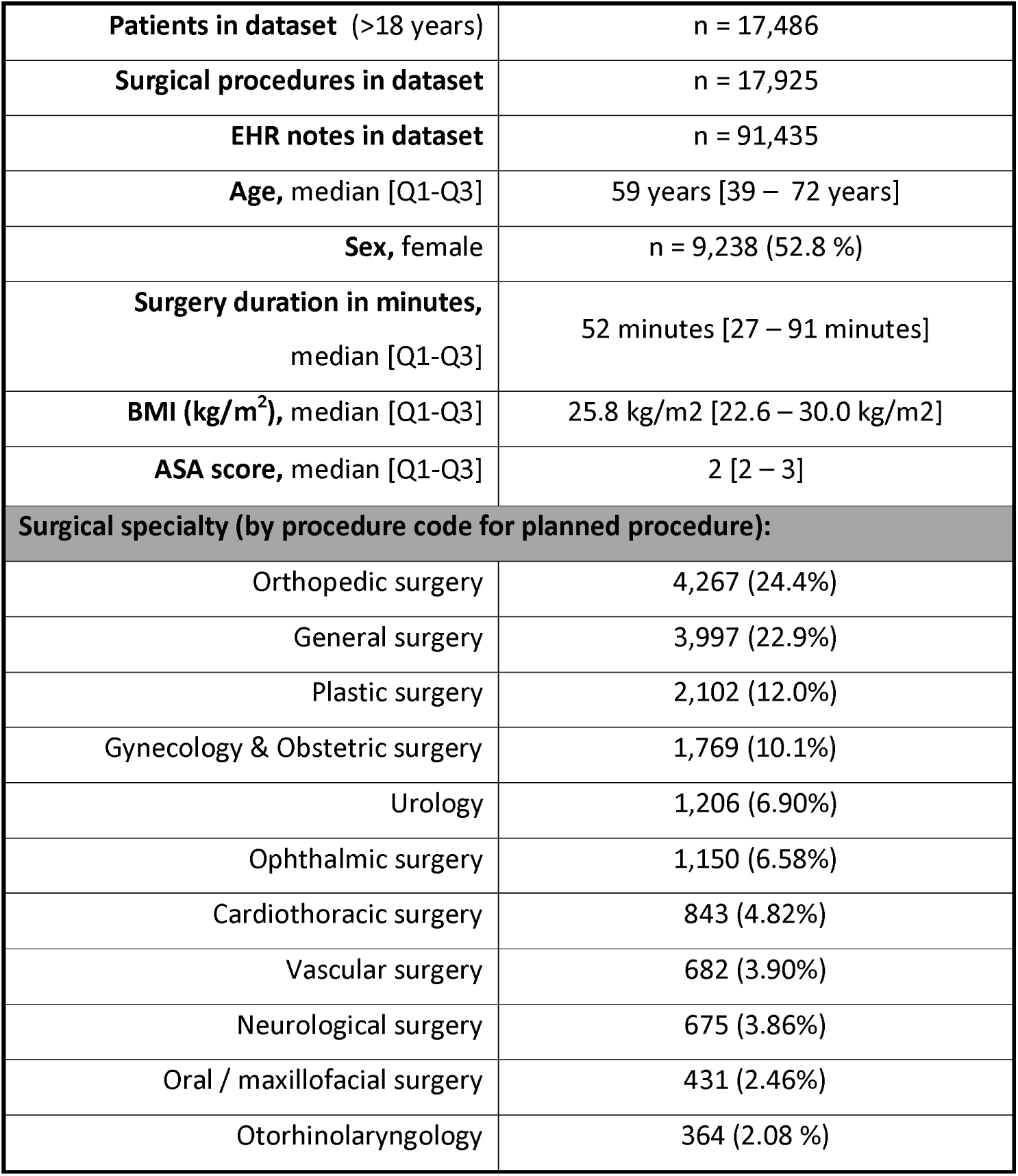
Demographics of the total dataset.

Table 2 presents the incidence of the eleven PCs in the validation and test sets, as identified by both ICD-10 tracking and manual curation. Manual curation consistently detected significantly more PCs than ICD-10 code tracking, with overall incidence rates over ten times higher. The incidence of complications by manual curation was 11.1% in the validation set and 13.5% in the test set. For ICD-10 coded PCs, only around one third were confirmed as actual complications by manual review (presented in the column “matching”). Specifically, 36.0% of the ICD-10 coded PCs in the validation set and 26.1% of the ICD-10 coded PCs in the test set were confirmed.

**Table 2:**
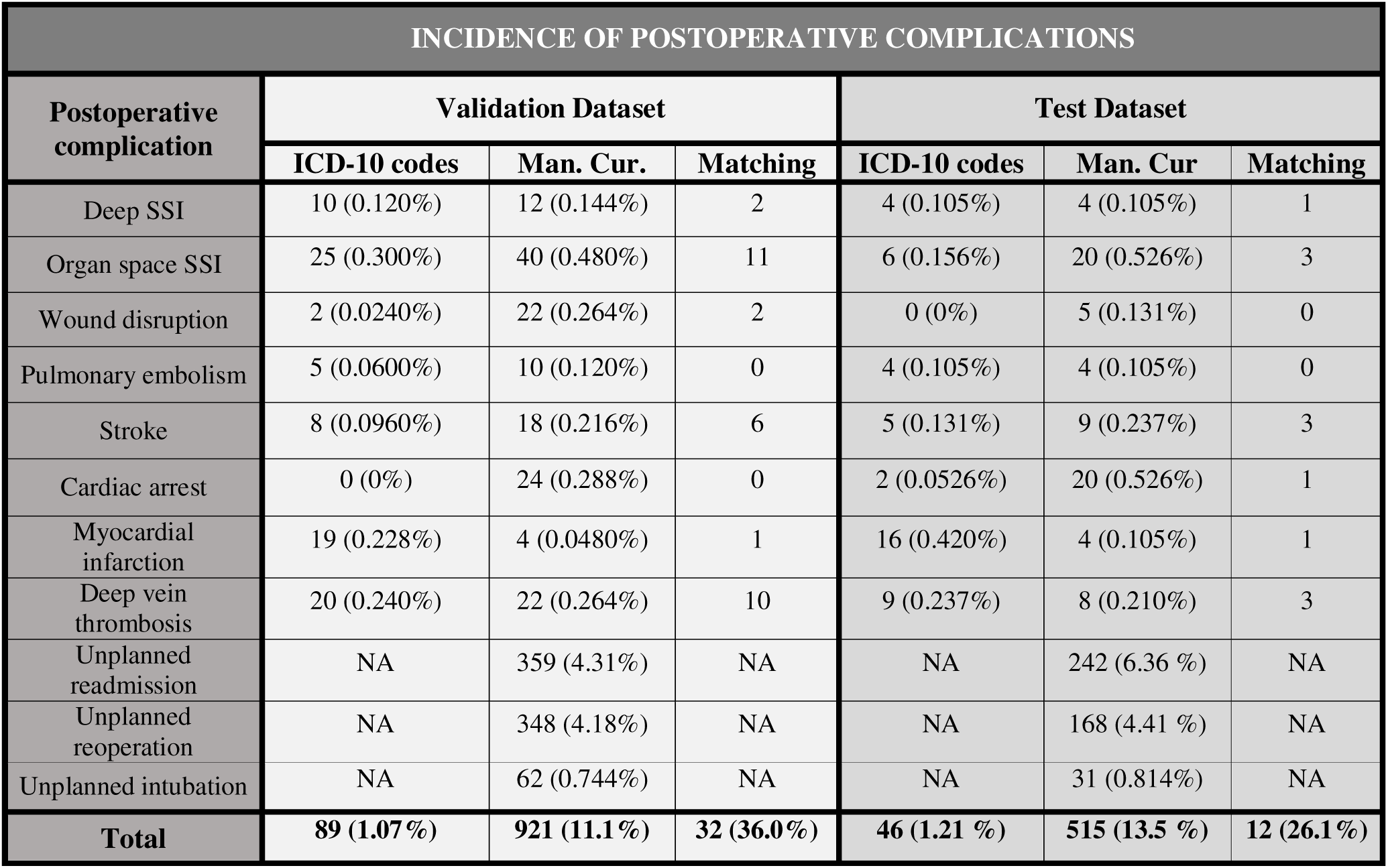
Incidence of the studied complications from the validation and test data sets determined by both ICD-10 codes and manual abstraction. Each subset includes a "Matching" column, which represents the cases where an ICD-10 coded complication was confirmed through manual review and was thus classified as the given PC by both ICD-10 and manual curation. The percentages in this column indicate the proportion of ICD-10 coded complications that were verified as complications upon manual review

Table 3 presents performance metrics for both the NLP algorithms and ICD-10 tracking compared to manual curation, which served as gold standard. Performance metrics are shown for both the SAM and the Human-In-The-Loop (HITL) approaches, as well as the thresholds.

**Table 3:**
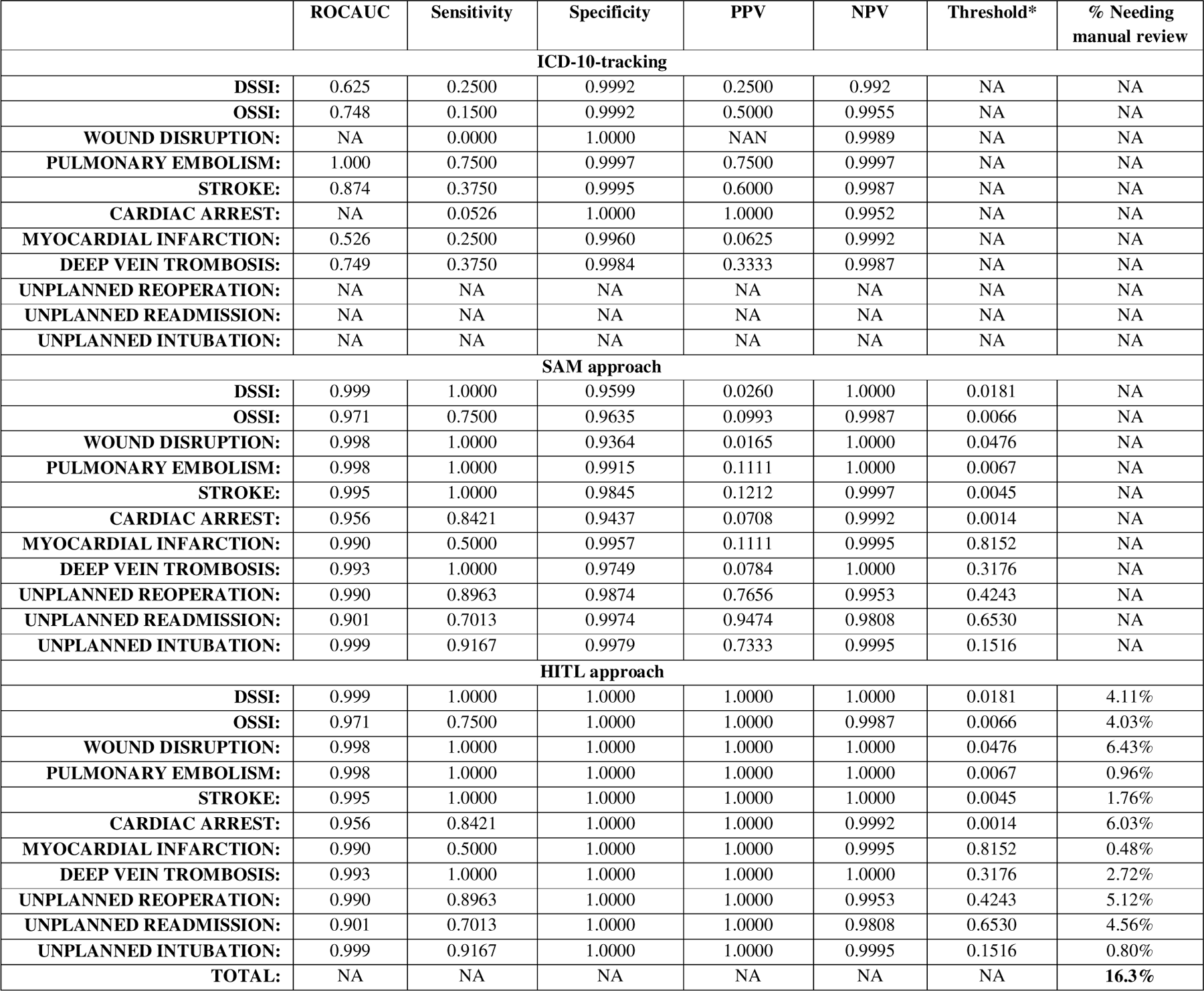
Performance metrics of the three different methods of identifying PCs. The Stand-Alone Model approach (SAM) represents the NLP algorithms’ performance by themselves with no human interaction. The Human In The Loop approach (HITL) represents the performance of the NLP algorithms’ in combination with human review, where high-probability cases identified by the NLP models were forwarded to human review. In this case it is presumed that manual review is the gold standard.

For ICD-10 tracking, ROC AUC values ranged from 0.531 (myocardial infarction) to 0.875 (pulmonary embolism). Wound disruption and cardiac arrest had an insufficient number of ICD-10 codes for ROC AUC calculations, and therefore these results are listed as NA. Sensitivity values ranged from 0.00 (wound disruption) to 0.375 (stroke and deep vein thrombosis), except for pulmonary embolism, which had a sensitivity of 0.750. Specificity values ranged between 0.996 and 1.00, PPV from 0.0625 to 1.00, and NPV from 0.992 to 0.999.

For the SAM approach, ROC AUC values ranged from 0.901 to 0.999, with sensitivity values from 0.701 to 1.00, except for myocardial infarction (0.500). Specificity ranged from 0.936 to 0.997, PPV from 0.0165 to 0.947, and NPV from 0.995 to 1.00.

In the HITL approach, ROC AUC, sensitivity, and NPV were identical to those in the SAM approach. However, due to human review being incorporated in this approach, specificity and PPV values were 1.00 across all PCs, as flagged cases were assumed correctly identified by the reviewer.

Comparative bar graphs (Figure 4) illustrate the methods for PC identification (ICD-tracking vs. SAM vs. manual curation). PCs with no ICD-10 codes available, SAM vs. manual curation is shown in Figure 5. True positives and false positives were presented for each PC as well.

**Figure 4:**
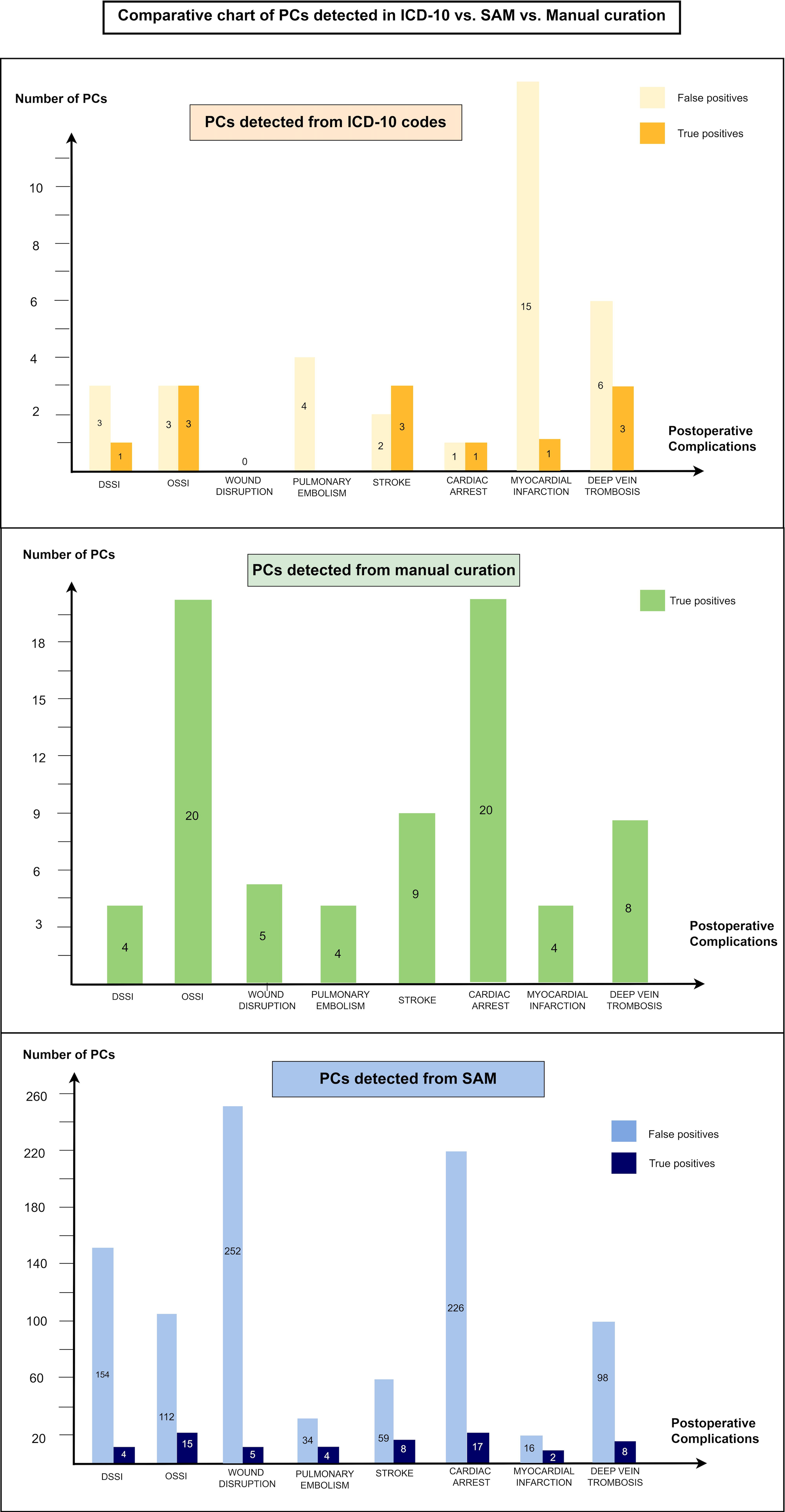
Comparative bar graphs of number of postoperative complications (PC) identified by ICD-10 codes, manual curation, and the “stand alone-model” (SAM) model for the test dataset. For each PC the number of both true positives and false positives are shown. DSSI: Deep Surgical Site Infections, OSSI: Organ / Space Surgical Site Infections

**Figure 5:**
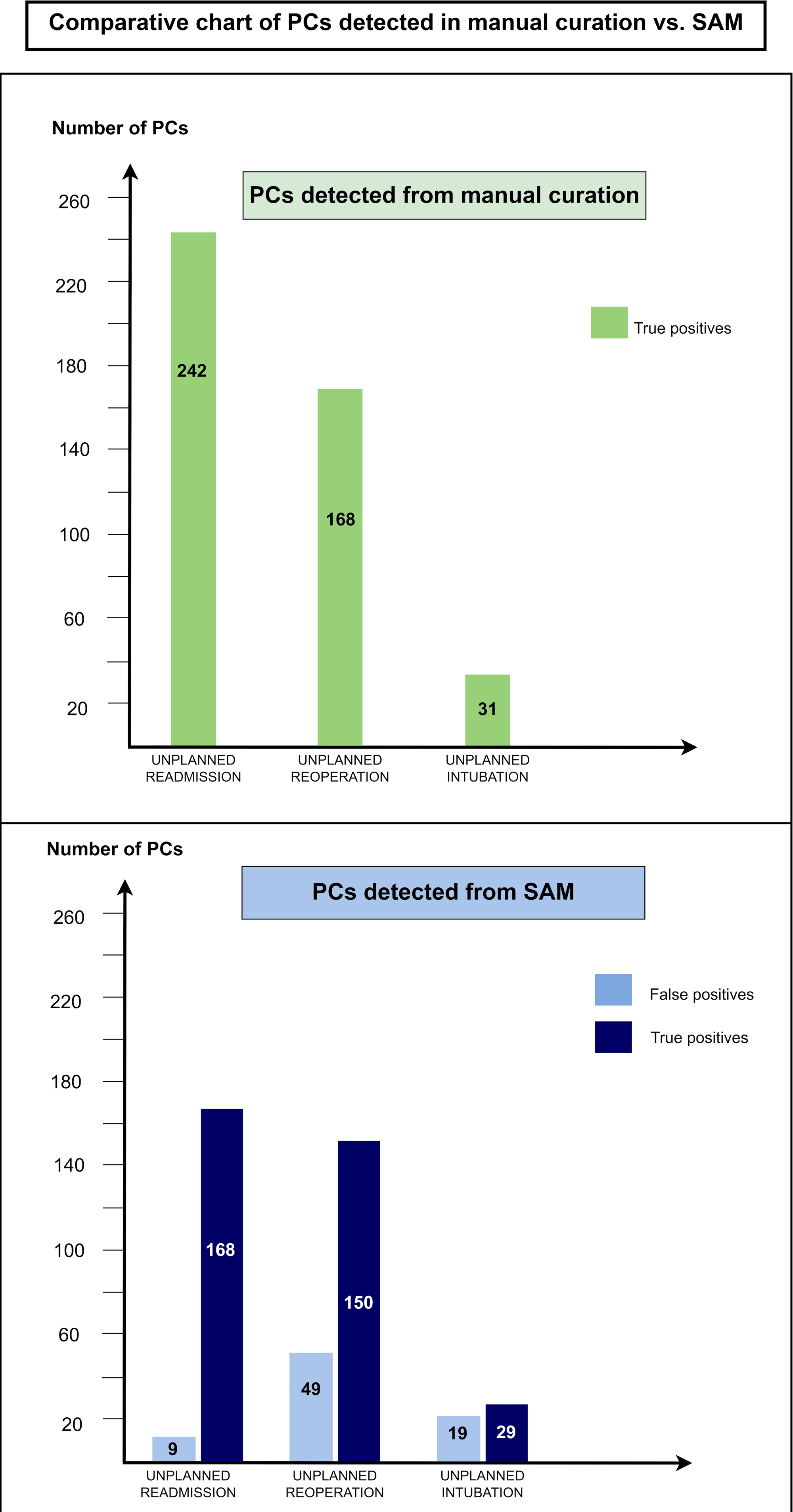
Comparative bar graphs displaying the number of postoperative complications (PC) identified by manual curation and the "stand-alone model" (SAM) for the test dataset only, as no ICD-10 codes for these PCs are offered. Each PC is represented by the number of true positives and false positive

Using the HITL-approach, between 0.48% (myocardial infarction) and 6.43% (wound disruption) of cases required manual review to achieve listed performance metrics. To detect all eleven PCs, 16.3% of all cases needing manual review.

## DISCUSSION

This study validated the use of NLP algorithms to detect a broad spectrum of postoperative complications. While the NLP models independently outperformed ICD-10 codes in sensitivity, the fully automated approach requires refinement due to a relatively high rate of false positives. We found that incorporating minimal human interaction produced a more accurate hybrid system (HITL), where only 16.3% of cases required manual review to achieve high sensitivity, specificity,

PPV and NPV. This reduced the manual workload by over 80% compared to fully manual quality improvement program, such as ACS NSQIP. Our findings suggest that when using NLP classification on unbalanced datasets (i.e., low incidence PCs), relying on ROC AUC alone may misrepresent model performance, and NLP remains insufficient for completely autonomous classification without sacrificing key quality metrics. This is in line with findings from previous studies.(6,18)

Even though prior studies have examined NLP for PC detection, most focus on a single complication, often surgical site infections(18–22). In comparison, our study evaluates a broad range of PCs. Previous studies report ROC AUC values between 0.855 and 0.921, with sensitivities ranging from 0.580 to 0.849. In contrast, our results show ROC AUC values ranging from 0.901 to 0.999, indicating superior performance, with the previously mentioned limitation in mind. Our sensitivities, ranging from 0.500 to 1.000 with an average of 0.873, are comparable to those reported in the literature(18–22). Other studies assessing NLP for multiple PCs report similar trends, with sensitivities from 0.560 and 0.950 for nine PCs(23), while another study achieved an average sensitivity of 0.830 for detecting 18 PCs(24). A 2021 systematic review and meta-analysis further that NLP models tend to outperform non-NLP models in sensitivity(25).

This study offers several strengths. Compared to the 45 studies included in the meta-analysis(25), our dataset is among the largest. Moreover, using a diverse, multi-specialty dataset increases robustness and generalizability. Additionally, comparing NLP with both the gold standard for PC- detection (manual curation) and the widely used ICD-10 codes tracking provided a comprehensive, multidimensional perspective on quality improvement initiatives. Furthermore, by covering a wide spectrum of PCs, the study offers a more realistic reflection of the diverse clinical setting relevant for future quality improvement strategies.

However, this study has limitations. First, not all EHR data types were included (e.g., imaging, nurse chart entries, microbiology results), which may have limited the Stand-Alone Model’s performance.

The quality of ICD-10 codes, which we based our training set on(18), presents another limitation. Recent findings from our research group indicate that ICD-10 codes not only underreport, but also misreport PCs. Using possibly low-quality ICD-10 codes for training the NLP-models may not be optimal.

The use of NLP and machine learning have key limitations. Although our models were trained and tested on multi-hospital data, their generalizability to other settings with different patient populations, workflows, or documentation styles remain undetermined. Dataset biases (e.g., age, gender, or race), may also affect model performance and potentially result in reduced accuracy for underrepresented subgroups.

To maximize the detection of true PCs, we optimized for sensitivity. Yet, this came at the expense of a lower positive predictive value (PPV), leading to a higher rate of false positive cases. As a result, the models cannot function autonomously. Therefore, we implemented the Human-In-The-Loop approach with manual review for high-probability cases. This ensured greater accuracy but limits the models’ efficiency.

Lastly, the “black box” nature of machine learning models (including NLP models), is a notable limitation. The decision-making process of these models, can lack transparency, possibly impacting trust and willingness to implement AI-tools in healthcare settings.

In summary, this study establishes NLP algorithms as an effective resource for detecting a wide range of postoperative complications from free medical text, achieving performance levels close near human curation and significantly outperforming ICD-10 tracking methods. These results support the potential of NLP to reduced time and labor associated with fully manual quality improvement programs. This approach opens the possibility of organizing a real-time, automated system for detecting, monitoring, and reducing postoperative complications on an international scale. Future studies should aim to further improve model performance and validate generalizability across diverse healthcare settings.

## Data Availability

Data availability statement: Due to GDPR-regulations and as this study utilizes patient sensitive data, the authors are not permitted to share the data without authorization. The dataset can be accessed with authorization from the Danish Patients Safety Board (Styrelsen for Patientsikkerhed) and the Danish Capital Region Data Safety Board (Videnscenter for Dataanmeldelser).

## Funding

Supported by a grant (#NNF19OC0055183) from the Novo Nordisk Foundation to MS

## Submission category

Original article

## Conflicts of interest

MS, AB and AT have founded Aiomic, a company developing AI models for healthcare systems. This study incorporates methodological scientific validations of the Aiomic product and underlying intellectual property, but the specific models within the study are not used for commercial purposes.

## Data availability statement

Due to GDPR-regulations and as this study utilizes patient sensitive data, the authors are not permitted to share the data without authorization. The dataset can be accessed with authorization from the Danish Patients Safety Board (Styrelsen for Patientsikkerhed) and the Danish Capital Region Data Safety Board (Videnscenter for Dataanmeldelser).

## Author contributions (CRediT)

EED, MS, AB and AT conceptualized the study and designed the methodology. EED and AB performed data curation and formal analysis. EED performed the collection of evidence, drafted the manuscript, and performed data visualization. MS supervised the study and, MS, AB, and AT reviewed and edited the final manuscript.

## REFERENCES

1. Ibrahim AM, Dimick JB. What Metrics Accurately Reflect Surgical Quality? Annu Rev Med. 2018 Jan 29;69:481–91.

2. Birkmeyer JD, Finks JF, O’Reilly A, Oerline M, Carlin AM, Nunn AR, et al. Surgical skill and complication rates after bariatric surgery. N Engl J Med. 2013 Oct 10;369(15):1434–42.

3. Chamely EA, Stulberg JJ. Measuring Quality at the Surgeon Level. Clin Colon Rectal Surg. 2023 Jul;36(4):233–9.

4. Stulberg JJ, Huang R, Kreutzer L, Ban K, Champagne BJ, Steele SR, et al. Association Between Surgeon Technical Skills and Patient Outcomes. JAMA Surg. 2020 Oct 1;155(10):960.

5. Dencker EE, Bonde A, Troelsen A, Sillesen M. ASSESSING THE ACCURACY GAP IN EARLY POSTOPERATIVE COMPLICATION SURVEILLANCE: ICD-10 CODES VERSUS MANUAL CURATION –CLINICAL AND ECONOMIC IMPLICATIONS. Scandinavian Journal of Surgery, accepted for publication 04-Oct-2024.

6. Dencker EE, Bonde A, Troelsen A, Sillesen M. Assessing the utility of natural language processing for detecting postoperative complications from free medical text. BJS Open. 2024 Mar 1;8(2).

7. Rodrigo-Rincon I, Martin-Vizcaino MP, Tirapu-Leon B, Zabalza-Lopez P, Abad-Vicente FJ, Merino-Peralta A. Validity of the clinical and administrative databases in detecting post-operative adverse events. Int J Qual Health Care. 2015 Aug;27(4):267–75.

8. Maass C, Kuske S, Lessing C, Schrappe M. Are administrative data valid when measuring patient safety in hospitals? A comparison of data collection methods using a chart review and administrative data. Int J Qual Health Care. 2015 Aug;27(4):305–13.

9. Lawson EH, Louie R, Zingmond DS, Brook RH, Hall BL, Han L, et al. A comparison of clinical registry versus administrative claims data for reporting of 30-day surgical complications. Ann Surg. 2012 Dec;256(6):973–81.

10. Romano PS, Chan BK, Schembri ME, Rainwater JA. Can Administrative Data Be Used to Compare Postoperative Complication Rates Across Hospitals? Med Care. 2002 Oct;40(10):856–67.

11. Richardson A, Pang T, Hitos K, Toh JWT, Johnston E, Morgan G, et al. Comparison of administrative data and the American College of Surgeons National Surgical Quality Improvement Program data in a New South Wales Hospital. ANZ J Surg. 2020 May;90(5):734–9.

12. Koch CG, Li L, Hixson E, Tang A, Phillips S, Henderson JM. What are the real rates of postoperative complications: elucidating inconsistencies between administrative and clinical data sources. J Am Coll Surg. 2012 May;214(5):798–805.

13. Henry LR, Minarich MJ, Griffin R, von Holzen UW, Hardy AN, Fornalik H, et al. Physician derived versus administrative data in identifying surgical complications. Fact versus Fiction. Am J Surg. 2019 Mar;217(3):447–51.

14. Hollenbeak CS, Boltz MM, Wang L, Schubart J, Ortenzi G, Zhu J, et al. Cost-effectiveness of the National Surgical Quality Improvement Program. Ann Surg. 2011 Oct;254(4):619–24.

15. Epelboym I, Gawlas I, Lee JA, Schrope B, Chabot JA, Allendorf JD. Limitations of ACS-NSQIP in reporting complications for patients undergoing pancreatectomy: underscoring the need for a pancreas-specific module. World J Surg. 2014 Jun;38(6):1461–7.

16. Ingraham AM, Richards KE, Hall BL, Ko CY. Quality Improvement in Surgery: the American College of Surgeons National Surgical Quality Improvement Program Approach. Adv Surg. 2010 Sep;44(1):251– 67.

17. Statistics Denmark [Internet]. [cited 2024 Nov 15]. Available from: https://www.dst.dk/da/Statistik/emner/borgere/befolkning/befolkningstal

18. Bonde A, Lorenzen S, Brixen G, Troelsen A, Sillesen M. Assessing the utility of deep neural networks in detecting superficial surgical site infections from free text electronic health record data. Front Digit Health. 2023;5:1249835.

19. Shi J, Lui S, Puiitt LCC, Luppens CL, Ferraro JP, Gundlapalli A V, et al. Using Natural Language Processing to improve EHR Structured Data-based Surgical Site Infection Surveillance. AMIA Annu Symp Proc . 2020;

20. Shi J, Hurdle JF, Johnson SA, Ferraro JP, Skarda DE, Finlayson SRG, et al. Natural language processing for the surveillance of postoperative venous thromboembolism. Surgery. 2021 Oct;170(4):1175–82.

21. Wu G, Cheligeer C, Southern DA, Martin EA, Xu Y, Leal J, et al. Development of machine learning models for the detection of surgical site infections following total hip and knee arthroplasty: a multicenter cohort study. Antimicrob Resist Infect Control. 2023 Sep 2;12(1):88.

22. Zhu Y, Simon GJ, Wick EC, Abe-Jones Y, Najafi N, Sheka A, et al. Applying Machine Learning Across Sites: External Validation of a Surgical Site Infection Detection Algorithm. J Am Coll Surg. 2021 Jun;232(6):963–971.e1.

23. FitzHenry F, Murff HJ, Matheny ME, Gentry N, Fielstein EM, Brown SH, et al. Exploring the Frontier of Electronic Health Record Surveillance. Med Care. 2013 Jun;51(6):509–16.

24. Bronsert M, Singh AB, Henderson WG, Hammermeister K, Meguid RA, Colborn KL. Identification of postoperative complications using electronic health record data and machine learning. Am J Surg. 2020 Jul;220(1):114–9.

25. Mellia JA, Basta MN, Toyoda Y, Othman S, Elfanagely O, Morris MP, et al. Natural Language Processing in Surgery: A Systematic Review and Meta-analysis. Ann Surg. 2021 May 1;273(5):900–8.

