## Supplementary data for "Validation of Natural Language Processing for Surgical Complication Surveillance: Detecting Eleven Postoperative Complications from Electronic Health Records"

Supplementary Material

**Table S1:** ICD-10 codes identifying the studied complication used to define structured administrative data.

| **DSSI** | |
| --- | --- |
| **Definition** | **ICD-10 code** |
| Infection following a procedure, deep | T81.4H |
| **OSSI** | |
| **Definition** | **ICD-10 code** |
| Anastomotic dehiscence / rupture | T81.3A |
| Postprocedural intra-abdominal abscess | T81.4B |
| Postprocedural subphrenic abscess | T81.4C |
| Postprocedural intra-abdominal infection | T81.4I |
| Postprocedural retroperitonal infection | T81.4J |
| **WOUND DISRUPTION** | |
| **Definition** | **ICD-10 code** |
| Disruption of operation wound, not elsewhere classified | T81.3 |
| **PULMONARY EMBOLISM** | |
| **Definition** | **ICD-10 code** |
| Postprocedural pulmonary embolism | T81.7D |
| Pulmonary embolism | I26 |
| Puerperal (pulmonary) embolism NOS | O88.2D |
| Obstetric pulmonary embolism NOS | O88.2E |
| **STROKE/CVA** | |
| **Definition** | **ICD-10 code** |
| Postprocedural cerebral apoplexy | T81.7Y1 |
| Intracerebral haemorrhage | I61 |
| Cerebral infarction | I63 |
| **CARDIAC ARREST** | |
| **Definition** | **ICD-10 code** |
| Cardiac arrest with successful resuscitation | I46.0 |
| Sudden cardiac death | I46.1 |
| **MYOCARDIAL INFARCTION** | |
| **Definition** | **ICD-10 code** |
| Postprocedural acute myocardial infarction | T81.7Y2 |
| Acute myocardial infarction | I21 |
| **DEEP VEIN TROMBOSIS** | |
| **Definition** | **ICD-10 code** |
| Vascular complications following a procedure, not elsewhere classified | T81.7C |
| Phlebitis and thrombophlebitis | I80 |

**Table S2:** The ACS-NSQIP 2018 Postoperative Complications definitions applied for manual abstraction.

### Deep incisional surgical site infection

**NSQIP definition:**

**Intent of Variable:** To capture the occurrence of infection that does not meet the criteria of Superficial Incisional SSI or Organ/Space SSI. These infections are typically more severe than the superficial SSI category.

**Definition:** Deep Incisional SSI is an infection which involves deep soft tissues. Deep soft tissues are typically any tissue beneath skin and immediate subcutaneous fat, for example fascial and muscle layers.

**Criteria:** An infection that occurs at the surgical site within 30 days after the primary procedure AND involves deep soft tissues AND at least ONE of the following:

- Purulent drainage from the deep incision but not from the organ/space component of the surgical site
- A deep incision spontaneously dehisces or is deliberately opened by a surgeon when the patient has at least one of the following signs or symptoms: fever (> 38⁰ C), localized pain, or tenderness, unless the site is culture-negative
- An abscess or other evidence of infection involving the deep incision is found on direct examination, during reoperation, or by histopathologic or radiologic examination
- Diagnosis of a deep incision SSI by a surgeon or attending physician

**Scenarios to clarify (Assign Variable):**

- Report an infection that involves both superficial and deep incision sites as Deep Incisional SSI Diagnosis of vaginitis in association with purulent drainage (i.e., from the cuff) after vaginal surgery
- Diagnosis of pharyngitis in association with purulent drainage after oral surgery

**Scenarios to clarify (Do Not Assign Variable):**

- Refer to superficial SSI variable for further clarification regarding superficial vaginitis and pharyngitis

**Notes:**

Only an SSI at the incision site of the primary procedure should only be assessed. Incision sites for “other” or “concurrent” procedures, if they are in distinctly different anatomical sites should not be assessed. If there is question as to whether or not an incision site was an integral portion of the primary procedure, include this site in your SSI assessment.

Criteria will be assigned when modifying terms such as “possible”, “probable”, “evolving”, “highly suspicious” or “suggestive” are used to describe an infection, in conjunction with otherwise meeting criteria. An SSI can only be assigned at or below the level of closure

Can be assigned multiple times within the 30-day postop period each time criteria is met.

### Organ-space surgical site infection

**NSQIP definition:**

**Intent of Variable:** To capture the occurrence of infection that does not meet the criteria of Superficial Incisional SSI or Deep Incisional SSI. This category of infection is typically the most severe and is more likely to require procedural intervention.

**Definition:** Organ/Space SSI is an infection that involves any part of the anatomy (e.g., organs or spaces), other than the incision, which was opened or manipulated during an operation.

**Criteria:** An infection that occurs within 30 days after the primary procedure AND involves any of the anatomy (e.g., organs or spaces), other than the incision, which was opened or manipulated during the operation AND at least ONE of the following:

A. Purulent drainage from a drain that is placed through a stab wound into the organ/space.

B. Organisms isolated from an aseptically obtained culture of fluid or tissue in the organ/space

C. An abscess or other evidence of infection involving the organ/space that is found on direct examination, during reoperation, or by histopathologic or radiologic examination. (Please see note below)

NOTE: If criterion C is met, please also refer to Appendix H. Organ/Space SSI Algorithm posted to the Resource Portal, Program Resources Tab, ACS NSQIP Operations Manual, Appendix H for additional guidance in assigning an Organ/Space SSI to a case.

D. Diagnosis of an organ/space SSI by a surgeon or attending physician

Scenarios to clarify (Assign Variable):

- Anastomotic leaks involving the GI tract or which involve enteric contents
- Anastomotic leaks involving the GU tract which involve evidence of an active infection (e.g., elevated WBC/fever attributed to the leak, diagnosis by physician, collection or leak is culture positive)
- Injury to intestine (e.g., enterotomy, iatrogenic injury) which results in a postoperative leak of entericcontents into the abdomen

Scenarios to clarify (Do Not Assign Variable):

- Fistulas alone, unless they independently meet the other criteria listed above
- Anastomotic leaks involving vasculature (e.g., lower extremity bypass), unless one of the 4 criteria above is met anastomotic leaks involving the GU tract, which do not meet criteria above
- A diagnosis of C diff in isolation (a positive culture without any other signs and symptoms of infection)

### Wound disruption

**NSQIP definition:**

**Intent of Variable:** To capture cases where the integrity of the surgical closure has been compromised.

**Definition:** The spontaneous reopening of a previously surgically closed wound.

**Criteria:** A spontaneous reopening of a surgically closed wound that occurs within 30 days after the primary procedure AND one of the following criteria A OR B below:

A. Abdominal site: refers primarily to loss of the integrity of fascial closure (or whatever closure was performed in the absence of fascial closure) OR

B. Other Surgical Sites: there must be a total breakdown of the surgical closure compromising the integrity of the procedure

**Scenarios to clarify (Assign Variable):**

- Tissue flap coverage where the surgical incisions, which were closed, have lost the integrity of closure. Above the knee amputation wound which spontaneously opens exposing the bone

**Scenarios to clarify (Do Not Assign Variable):**

- An ostomy with a small separation around it

**Notes**:

If a wound is closed in a subsequent procedure within the 30 day postop period, assign disruption based on this closure. Can be assigned multiple times within the 30 day postop period each time criteria is met.

### Pulmonary embolism

**NSQIP definition:**

**Intent of Variable:** The identification of a new blood clot in a pulmonary artery causing obstruction (complete or partial) of the blood supply to the lungs. However, since there are not always preoperative studies proving that a clot or thrombus was not present preoperatively, the technical specification of the variable requires only a “new diagnosis”- in other words the clot or thrombus was not previously known.

**Definition:** Lodging of a blood clot in the pulmonary artery with subsequent obstruction of blood supply to the lung parenchyma. The blood clots usually originate from the deep leg veins or the pelvic venous system.

**Criteria:** A pulmonary embolism must be noted during or within 30 days after the primary procedure AND the following criteria, A AND B below:

A. New diagnosis of a new blood clot in a pulmonary artery

AND

B. The patient has a V-Q scan interpreted as high probability of pulmonary embolism or a positive CT exam, TEE, pulmonary arteriogram, CT angiogram, or any other definitive imaging modality (including direct pathology examination such as autopsy)

**Scenarios to clarify (Assign Variable):**

**Scenarios to clarify (Do Not Assign Variable):**

- Pulmonary emboli diagnosed prior to the primary procedure
- Cement PE (if this diagnosis is definitive)
- Fat PE (if this diagnosis is definitive)

**Notes:**

Can be assigned multiple times within the 30-day postop period each time criteria is met.

### Stroke/CVA

**NSQIP definition:**

**Intent of Variable:** To identify patient(s) who developed an acute cerebral vascular accident or acute stroke during or after surgery affecting their physiology as described.

**Definition:** An interruption or severe reduction of blood supply to the brain resulting in severe dysfunction.

**Criteria:** A Stroke/Cerebral Vascular Accident (CVA) must be noted during or within 30 days after the primary procedure AND one of the following A OR B below:

A. There is motor, sensory, or cognitive dysfunction which persists for 24 hours or more in the setting of a suspected stroke.

OR

B. If a specific timeframe for the dysfunction is not documented in the medical record, but there is a diagnosis of a stroke, assign the occurrence, unless documentation specifically states that the motor, sensory, or cognitive dysfunction resolved within 24 hours.

**Notes:**

Can be assigned multiple times within the 30 day postop period each time criteria is met.

### Cardiac arrest requiring cardiopulmonary resuscitation

**NSQIP definition:**

**Intent of Variable:** To identify patient(s) who experienced a cardiac arrest or dysfunction and required the initiation of CPR.

**Definition:** The absence of cardiac rhythm or presence of a chaotic cardiac rhythm requiring the initiation of cardiopulmonary resuscitation.

**Criteria:** Cardiac Arrest Requiring CPR must be noted intraoperatively or within 30 days after the primary procedure AND one of the following three scenarios (A or B or C) below:

A. The absence of a cardiac rhythm or presence of chaotic cardiac rhythm requiring the initiation of chest compressions

OR

B. Patients in pulseless VT or V-Fib in which defibrillation is performed with or without chest compressions

OR

C. Patients with automatic implantable cardioverter defibrillator (AICD) that fires and the patient has loss of consciousness

**Scenarios to clarify (Assign Variable):**

- PEA (pulseless electrical activity) arrests requiring chest compressions
- Patient receives open cardiac massage
- Patient requires the initiation of chest compressions but documentation indicates that chest compressions were not performed due to DNR status

**Scenarios to clarify (Do Not Assign Variable):**

- Patient receives initial ACLS medications but do not proceed to the initiation of chest compressions (except for VT or V-Fib as noted above)

**Notes:**

Can be assigned multiple times within the 30 day postop period each time criteria is met.

### Myocardial infarction

**NSQIP definition:**

**Intent of Variable:** To identify patient(s) who sustain an acute myocardial infarction (intraop or postop) affecting their physiology as described.

**Definition:** Blockage of blood flow to the heart causing damage or death to part of the heart muscle.

**Criteria:** An acute myocardial infarction must be noted intraoperatively OR within 30 days after the primary procedure AND one of the following three scenarios (A or B or C) below:

A. Documentation of ECG changes indicative of acute MI (one or more of the following three):

1. ST elevation > 1 mm in two or more contiguous leads

2. New left bundle branch block

3. New q-wave in two or more contiguous leads

OR

B. New elevation in troponin greater than three times upper level of the reference range in the setting of suspected myocardial ischemia

OR

C. Physician, PA or NP diagnosis of myocardial infarction

**Scenarios to clarify (Assign Variable):**

- Diagnosis of demand ischemia along with any troponin elevation.

**Scenarios to clarify (Do Not Assign Variable):**

- A diagnosis of MI is rendered, however, cardiology is consulted and renders an official opinion that signs and symptoms are unrelated to an MI

**Notes**:

The article in which the ACS NSQIP MI criteria is based {Circulation. 2007; 116: 2634-2653} can be referenced at the following website:http://circ.ahajournals.org/content/116/22/2634

- Can be assigned multiple times within the 30 day postop period each time criteria is met.

Vein Thrombosis Requiring Therapy

**NSQIP definition:**

**Intent of Variable:** To identify patient(s) that developed a new blood clot or thrombus within the **venous system** postoperatively affecting their physiology and requiring treatment as described. However, since there are not always preoperative studies proving that a clot or thrombus was not present preoperatively, the technical specification of the variable requires only a “new diagnosis”- in other words, the clot or thrombus was not previously known.

**Definition:** New diagnosis of blood clot or thrombus within the **venous system** (superficial or deep) which may be coupled with inflammation and requires treatment.

**Criteria:** Must be noted within 30 days after the primary procedure **AND** one of the following A or B below:

A. New Diagnosis of a [new] venous thrombosis (superficial or deep), confirmed by a duplex, venogram, CT scan, or any other definitive imaging modality (including direct pathology examination such as autopsy) **AND** the patient **must be treated.** Treatment includes anticoagulation therapy, placement of a vena cava filter, clipping of the vena cava, or thrombectomy. If the record indicates that treatment was warranted but there was no additional appropriate treatment option available, this would meet treatment criteria.

**OR**

B. As per (A) above, but the patient or decision maker has refused treatment. There must be documentation in the medical record of the [patient’s] refusal of treatment.

**Scenarios to clarify (Assign Variable):**

- Internal jugular (IJ) clots
- Cephalic Vein clots
- Portal vein clots
- Patient requires therapy, but refuses
- Chronic venous thrombosis present preoperatively, which are also noted postoperatively with evidence of

new progression

**Scenarios to clarify (Do Not Assign Variable):**

- Chronic venous thrombosis present preoperatively, which are also noted postoperatively but without evidence of new progression
- If only an intravenous catheter is thrombosed and the vein is not.
- Arterial clots
- Aspirin alone does not constitute treatment sufficient to assign vein thrombosis requiring therapy

**Notes:**

- Can be assigned multiple times within the 30-day postop period each time criteria are met.

**Table S3:** Data preprocessing **–** Inclusion / exclusion criteria

- All cases between May 2016 and November 2021 are **included.**

*(With 30-day follow-up)*

- All cases with missing variables for “start time” or “end time” are **excluded**

- All cases with status not marked as “Completed” are **excluded**

- All cases with irrelevant or missing procedure codes (planned SKS code) are **excluded**

*(Only including prefixes: “KA”, “KB”, “KC”, “KD”, “KE”, “KF”, “KG”, “KH”, “KJ”, “KK”, “KL”, “KM”, “KN”, “KP”,“KQ”)*

- Only primary cases are **included.**

*(A case is primary if no other case exists for the same patient within 30 days before.)*

- All cases from May 2016 to Oct. 31^st^ with ICD-10 code for SSSI are **included** in **the training dataset.**
